# Effectiveness of COVID-19 Vaccination Schedules Against Severe COVID-19 in Children Aged 6 Months to 4 Years in Brazil: A Population-Based Cohort Study (2023–2024)

**DOI:** 10.64898/2026.03.03.26347530

**Authors:** Isaac Negretto Schrarstzhaupt, Fredi Alexander Diaz-Quijano

## Abstract

**Background:** Although the impact of COVID-19 vaccination is widely documented in the general population, the evidence on its effectiveness in children under 5 years of age is still limited. In this context, the continuation of vaccination programs in this age group has been debated globally. Consequently, we estimated the effectiveness of the 3-dose series of BNT162b2 (Pfizer-BioNTech) in children aged 6 months to 4 years and the complete 2-dose series of CoronaVac (Sinovac) in children aged 3 to 4 in reducing the risk of hospitalizations due to COVID-19-attributed severe acute respiratory infection (SARI) in Brazil.

**Methods:** We conducted a retrospective cohort study in 24 Brazilian municipalities, using surveillance data. We evaluated vaccine effectiveness in reducing the incidence rate of COVID-19-attributed SARI hospitalizations from July 2023 to December 2024. Covariate adjustments, defined a priori based on a conceptual model represented by a directed acyclic graph (DAG), were implemented using random-effects Poisson regression models. We also analyzed alternative vaccination schedules and obtained age-specific estimates of effectiveness.

**Results:** The cohort comprised 37.6 million person-months of follow-up and 1,384 COVID-19-attributed SARI hospitalizations, including 27 associated deaths. The 3-dose series of BNT162b2 vaccine had an effectiveness of 97% (IRR 0.03, 95%CI 0.01-0.10) in the group aged 6 months to 4 years, with no significant differences among age-specific estimates. No deaths occurred among children who completed the 3-dose series, whereas four deaths were observed among those with fewer doses. The effectiveness of CoronaVac was small and not statistically significant (IRR 0.96, 95%CI 0.57-1.62). However, no deaths were recorded among children who received any number of CoronaVac doses, and no COVID-19-attributed SARI hospitalizations were observed among those who received a third dose of this vaccine.

**Conclusions:** The 3-dose series of the mRNA vaccine (BNT162b2) had high and consistent effectiveness in protecting against severe COVID-19 in children aged 6 months to 4 years. These findings support the maintenance of routine COVID-19 vaccination in this age group.

## INTRODUCTION

COVID-19 had numerous health impacts,^1–4^ including a considerable number of severe acute respiratory infection (SARI) hospitalizations.^5^ Vaccines substantially reduced severe cases and deaths;^6–8^ however, children were among the last groups to be vaccinated due to their lower perceived risk.^9,10^ As adult immunity increased, severe cases and deaths became proportionally concentrated in vulnerable and under-vaccinated populations, increasing the relative contribution of children to severe cases and deaths.^11–13^

In Brazil, pediatric COVID-19 vaccination was introduced in 2022, coinciding with the circulation of the Omicron variant and its sublineages.^14^ The Brazilian National Immunization Program (PNI) initially recommended the CoronaVac (Sinovac) vaccine for children aged 3 years and older (July 2022),^15^ followed by the BNT162b2 (Pfizer-BioNTech) vaccine for children aged 6 months to 4 years old (December 2022).^14^ However, despite COVID-19 vaccination campaigns, vaccine coverage in these age groups remained low.^16^

Although clinical trials demonstrated safety and efficacy^10^ and there are effectiveness estimates in some countries,^17,18^ evidence in children under 5 years of age, especially in the context of the Omicron variant, remains limited. Moreover, several countries have reconsidered the COVID-19 vaccine recommendations for this age group.^19–22^ Given this scenario, evaluating vaccine impact on clinically relevant outcomes is essential to inform immunization policy.

This study aimed to estimate the effectiveness of the COVID-19 vaccination series recommended for children in Brazil, including the 3-dose series of BNT162b2 for children aged 6 months to 4 years and the 2-dose series of CoronaVac for children aged 3 and 4 years, in reducing COVID-19-attributed SARI rates. Additionally, we evaluated schedules with different numbers of doses and obtained age-specific estimates of effectiveness.

## METHODS

### Study design and period

This population-based retrospective cohort study used surveillance data, covering children aged 6 months to 4 years, residing in Brazilian municipalities. We aimed to avoid issues arising from sparse data, ensuring enough exposed individuals and events for the analysis. Thus, we chose the 24 municipalities with the most events among the 200 municipalities with the highest coverage (Figure 1).

**Figure 1:**
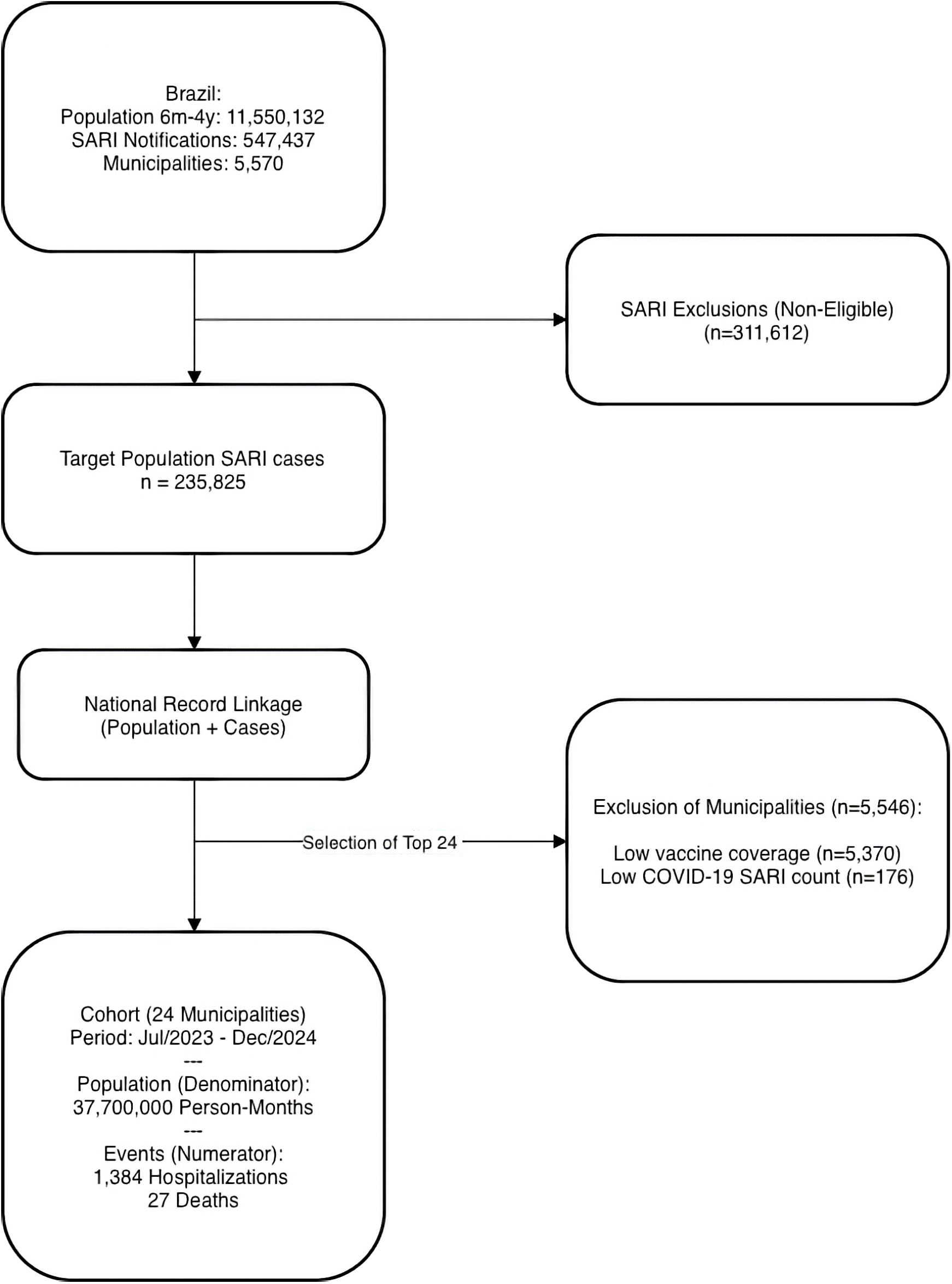
Flowchart for the database used in the study.

As this study utilized publicly available and completely anonymized surveillance data, the requirement for institutional review board approval and written informed consent was waived in accordance with national ethical guidelines. The data analyses were planned by FADQ and conducted by INS.

### Data sources

We analyzed the SARI hospitalizations meeting clinical criteria (dyspnea, respiratory distress, hypotension, or O_2_ saturation <95%) and attributed to COVID-19 via laboratory (RT-PCR, RT-LAMP, or rapid antigen test) or clinical-epidemiological criteria, per Ministry of Health guidelines.^23^ Data came from the Influenza Epidemiological Surveillance Information System (SIVEP-Gripe), the mandatory registry for hospitalized SARI cases by any cause.^24,25^

Moreover, we used the official COVID-19 campaign vaccination records from the National Health Data Network (RNDS).^24,26,27^ The monthly number of unvaccinated children was calculated by subtracting cumulative vaccinated children from the total population, using single-year age estimates (monthly/annual) per municipality according to the 2022 Demographic Census projections of the Brazilian Institute of Geography and Statistics (IBGE).^28^ All data are publicly available and anonymized.

To calculate the monthly incidence rates, the population on the first day of each month was used as the denominator for that month, assuming it approximated the average population at risk, a premise supported by analysis of monthly vaccine coverage variation. Because the first half of 2023 corresponded predominantly to the vaccine introduction period, monthly variations in the denominators exceeded 6%, potentially compromising this assumption (Supplemental Table S1). From July 2023 onwards, the variation was <3%. Therefore, we selected the period from July 1, 2023, to December 31, 2024, to minimize bias due to substantial within-month variation in the population at risk.

### Data Integration and Vaccine Identification

Lacking a unique and universal individual identifier in the public databases, we linked numerator and denominator data using aggregated strata (municipality of residence, age in months, and calendar month). Vaccines were identified by the Ministry of Health immunobiological codes:^16,29^

- BNT162b2: 87, 99, 102 and 103 (We grouped all manufacturer codes).
- CoronaVac: 86 and 98.

The dataset processed in this study, which came from the RNDS, covers 95% of the total doses on the official vaccination dashboard of the Ministry of Health, including 100% of the CoronaVac doses and 97% of the BNT162b2 doses.^16^ Although the mRNA-1273 (Moderna) vaccine was identified, it was not evaluated as the exposure of interest due to its late introduction into the schedule, but it was considered in the statistical models as an adjustment covariate (see below).

### Estimands and statistical analysis

We estimated BNT162b2 effectiveness in the population aged 6 months to 4 years, and CoronaVac effectiveness in the population aged 3 to 4 years, in reducing the incidence rate of COVID-19-attributed SARI during the study period. These age groups correspond to the approved indications for use by the Ministry of Health.^15^ The unit of analysis was defined as the set of children of a specific age, in a given month of follow-up, municipality, and vaccination status category.

In each unit, the number of incident COVID-19-attributed SARI hospitalizations occurring in that stratum (numerator) and the total child-months (rate denominator) were aggregated. Children could contribute person-time to multiple consecutive months, according to their age, municipality, and vaccination status; however, each SARI episode was counted only once and attributed to the unit defined by the month and vaccination status at the time of occurrence. Doses received in the 15 days before hospitalization were disregarded when defining the vaccination status. Given that the denominators correspond to the population at risk at the beginning of each month (approximately 15 days before the midpoint of the monthly period), this approach ensured temporal alignment between vaccination status classification in the population at risk and in the corresponding outcomes.

For each estimate of interest, we used random-effects Poisson regression models, aggregated by municipality, to estimate incidence rate ratios (IRR). Initially, we estimated the overall effects, adjusting for covariates suggested by a conceptual model represented by a DAG (Supplemental Figure S1), including age, temporal trend, and use of other vaccines, as well as interaction terms between these covariates when statistically significant. We additionally included, as a contextual covariate, the municipality-level incidence of COVID-19–attributed SARI among children under 5 years in 2022, given its plausible association with both subsequent vaccination coverage and the risk of the outcome during the study period.^30^

Each vaccine of interest was evaluated as an independent exposure, with the others included as covariates, and interactions arising from dose combinations between the vaccines were also tested in multivariable models. Next, we evaluated interaction terms between the vaccine of interest and the age and time variables, as potential effect modifiers.

Age-specific estimates were also obtained. In strata where no events were recorded in the vaccinated group, we presented crude estimates, with confidence intervals calculated by the exact Poisson method. Finally, when the vaccine was associated with a statistically significant reduction in the COVID-19-attributed SARI rate, we reported the vaccine effectiveness (VE), calculated as:

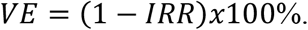

Two sensitivity analyses were conducted to evaluate the data robustness. The exclusion of the São Paulo municipality from the study, due to the potential selection bias from the migration of the local information system (Vacivida) to the RNDS,^31^ did not alter the direction or the statistical significance of the observed associations (Supplemental Table S3). Additionally, the denominator validation, comparing the 2022 Census with the live birth records (SINASC 2019-2024),^32^ resulted in a difference of 2%, indicating high agreement between the datasets (Supplemental Table S4).

## RESULTS

The analyzed cohort accumulated 37,687,410 person-months of observation across the 24 selected municipalities. There were 1,384 COVID-19-attributed SARI hospitalization events, resulting in an overall incidence of 3.67 cases per 100,000 person-months, including 27 associated deaths.

Regarding the BNT162b2 vaccine, the unvaccinated category accounted for most of the follow-up time (28 million person-months), 83% of hospitalizations (1,175), and 23 of the 27 deaths. There were no deaths recorded in children with the complete primary series (3 doses). Of the 4 deaths that occurred among the vaccinated, 2 were in the 1-dose and 2 in the 2-dose categories (Table 1).

**Table 1:** Observation time and events by age group and BNT162b2 vaccination status (July 2023-December 2024)

| Age group and vaccination status | Person-months | COVID-19-attributed SARI | Incidence (per 100,000 person-months) | Deaths |
| --- | --- | --- | --- | --- |
| *6-11 months* |  |  |  |  |
| Unvaccinated | 3,427,416 | 560 | 16.33 | 13 |
| BNT162b2, 1 dose | 210,621 | 16 | 7.60 | 0 |
| BNT162b2, 2 doses | 122,606 | 7 | 5.71 | 0 |
| BNT162b2, 3 doses | 28,233 | 0 | 0 | 0 |
| **Subtotal** | **3,788,876** | **583** | **15.38** | **13** |
| *1 year* |  |  |  |  |
| Unvaccinated | 5,572,891 | 331 | 5.94 | 9 |
| BNT162b2, 1 dose | 695,314 | 34 | 4.89 | 0 |
| BNT162b2, 2 doses | 622,766 | 48 | 7.71 | 0 |
| BNT162b2, 3 doses | 585,566 | 1 | 0.17 | 0 |
| **Subtotal** | **7,476,537** | **414** | **5.54** | **9** |
| *2 years* |  |  |  |  |
| Unvaccinated | 5,976,247 | 110 | 1.84 | 1 |
| BNT162b2, 1 dose | 881,863 | 12 | 1.36 | 0 |
| BNT162b2, 2 doses | 737,556 | 30 | 4.07 | 1 |
| BNT162b2, 3 doses | 781,783 | 2 | 0.25 | 0 |
| **Subtotal** | **8,377,449** | **154** | **1.84** | **2** |
| *3 years* |  |  |  |  |
| Unvaccinated | 6,285,258 | 85 | 1.35 | 0 |
| BNT162b2, 1 dose | 1,059,433 | 10 | 0.94 | 1 |
| BNT162b2, 2 doses | 793,259 | 28 | 3.53 | 1 |
| BNT162b2, 3 doses | 748,124 | 0 | 0 | 0 |
| **Subtotal** | **8,866,074** | **123** | **1.39** | **2** |
| *4 years* |  |  |  |  |
| Unvaccinated | 6,778,621 | 89 | 1.31 | 0 |
| BNT162b2, 1 dose | 1,299,644 | 12 | 0.92 | 1 |
| BNT162b2, 2 doses | 622,184 | 9 | 1.45 | 0 |
| BNT162b2, 3 doses | 453,392 | 0 | 0 | 0 |
| **Subtotal** | **9,153,841** | **110** | **1.20** | **1** |
| *6 months-4 years* |  |  |  |  |
| Unvaccinated | 28,040,433 | 1,175 | 4.19 | 23 |
| BNT162b2, 1 dose | 4,146,875 | 84 | 2.02 | 2 |
| BNT162b2, 2 doses | 2,898,371 | 122 | 4.21 | 2 |
| BNT162b2, 3 doses | 2,597,098 | 3 | 0.12 | 0 |
| **Total analyzed** | **37,682,777** | **1,384** | **3.67** | **27** |

Regarding CoronaVac, 92% of the children with the complete primary series were 4 years old. Although its indication was for children aged 3 years and older, it was administered to a small proportion of children aged 6 months to 2 years. There were no deaths in any of the vaccinated strata (Table 2).

**Table 2:** Observation time and events by age group and CoronaVac vaccination status (July 2023-December 2024)

| Age group and vaccination status | Person-months | COVID-19-attributed SARI | Incidence (per 100,000 person-months) | Deaths |
| --- | --- | --- | --- | --- |
| *6-11 months* |  |  |  |  |
| Unvaccinated | 3,788,734 | 583 | 15.39 | 13 |
| CoronaVac, 1 dose | 153 | 0 | 0 | 0 |
| CoronaVac, 2 doses | 19 | 0 | 0 | 0 |
| CoronaVac, 3 doses | 4 | 0 | 0 | 0 |
| **Subtotal** | **3,788,910** | **583** | **15.39** | **13** |
| *1 year* |  |  |  |  |
| Unvaccinated | 7,476,114 | 410 | 5.48 | 9 |
| CoronaVac, 1 dose | 1,247 | 4 | 320.77 | 0 |
| CoronaVac, 2 doses | 145 | 0 | 0 | 0 |
| CoronaVac, 3 doses | 0 | 0 | 0 | 0 |
| **Subtotal** | **7,477,506** | **414** | **5.54** | **9** |
| *2 years* |  |  |  |  |
| Unvaccinated | 8,375,836 | 154 | 1.84 | 2 |
| CoronaVac, 1 dose | 2,297 | 0 | 0 | 0 |
| CoronaVac, 2 doses | 712 | 0 | 0 | 0 |
| CoronaVac, 3 doses | 11 | 0 | 0 | 0 |
| **Subtotal** | **8,378,856** | **154** | **1.84** | **2** |
| *3 years* |  |  |  |  |
| Unvaccinated | 8,694,435 | 116 | 1.33 | 2 |
| CoronaVac, 1 dose | 103,491 | 2 | 1.93 | 0 |
| CoronaVac, 2 doses | 88,888 | 5 | 5.62 | 0 |
| CoronaVac, 3 doses | 470 | 0 | 0 | 0 |
| **Subtotal** | **8,887,284** | **123** | **1.38** | **2** |
| *4 years* |  |  |  |  |
| Unvaccinated | 7,329,864 | 87 | 1.19 | 1 |
| CoronaVac, 1 dose | 801,273 | 12 | 1.50 | 0 |
| CoronaVac, 2 doses | 1,017,357 | 11 | 1.18 | 0 |
| CoronaVac, 3 doses | 6,360 | 0 | 0 | 0 |
| **Subtotal** | **9,154,854** | **110** | **1.20** | **1** |
| *3-4 years* |  |  |  |  |
| Unvaccinated | 16,024,299 | 203 | 1.27 | 3 |
| CoronaVac, 1 dose | 904,764 | 14 | 1.55 | 0 |
| CoronaVac, 2 doses | 1,106,245 | 16 | 1.45 | 0 |
| CoronaVac, 3 doses | 6,830 | 0 | 0 | 0 |
| **Total 3-4 years** | **18,042,138** | **233** | **1.29** | **3** |
| **Total** | **37,687,410** | **1,384** | **3.67** | **27** |

### Effectiveness estimates

The complete 3-dose series of BNT162b2 was associated with a 97% reduction in the incidence rate of COVID-19-attributed SARI (IRR 0.03, 95% CI 0.01-0.10). In more than 2.5 million person-months corresponding to this series, there were only 3 recorded events. This effectiveness was not modified by time, age, or the administration of other vaccines (p >0.60 for all interaction terms), remaining consistent across all ages (Table 3).

**Table 3:** Incidence Rate Ratios of the BNT162b2 vaccine against COVID-19-attributed SARI: overall and age-specific estimates.

|  | Adjusted IRR (95 CI%)<sup>a</sup> | VE-% (95 CI%)<sup>c</sup> | p-value |
| --- | --- | --- | --- |
| **Overall estimate**<br>**(6 months-4 years)** |  |  |  |
| 1 dose | 0.62 (0.49 - 0.77) | 38 (23 - 51) | <0.001 |
| 2 doses | 1.15 (0.95 - 1.38) | - | 0.15 |
| 3 doses | 0.03 (0.01 - 0.10) | 97 (90 - 99) | <0.001 |
| **Age-specific estimate** |  |  |  |
| *6-11 months* |  |  |  |
| 1 dose | 0.43 (0.26 - 0.71) | 57 (29 - 74) | <0.001 |
| 2 doses | 0.30 (0.14 - 0.63) | 70 (37 - 86) | <0.001 |
| 3 doses | 0.00<sup>d</sup> (0.00 - 0.80) | 100 (20 - 100)<sup>d</sup> | 0.01 |
| *1 year* |  |  |  |
| 1 dose | 0.83 (0.59 - 1.19) | - | 0.32 |
| 2 doses | 1.26 (0.93 - 1.71) | - | 0.13 |
| 3 doses | 0.03 (0.00 - 0.19) | 97 (81 - 100) | <0.001 |
| *2 years* |  |  |  |
| 1 dose | 0.75 (0.41 - 1.37) | - | 0.35 |
| 2 doses | 2.26 (1.51 - 3.40) | - | <0.001 |
| 3 doses | 0.15 (0.04 - 0.60) | 85 (40 - 96) | 0.008 |
| *3 years* |  |  |  |
| 1 dose | 0.69 (0.36 - 1.34) | - | 0.27 |
| 2 doses | 2.74 (1.77 - 4.22) | - | <0.001 |
| 3 doses | 0.00<sup>d</sup> (0.00 - 0.37) | 100 (63-100)<sup>d</sup> | < 0.001 |
| *4 years* |  |  |  |
| 1 dose | 0.79 (0.44 - 1.50) | - | 0.47 |
| 2 doses | 1.36 (0.68 - 2.74) | - | 0.39 |
| 3 doses | 0.00<sup>d</sup> (0.00 - 0.63) | 100 (37-100)<sup>d</sup> | 0.003 |

Notes:
a) Adjusted for age, time and other vaccines
b) Adjusted for time and other vaccines.
c) $VE = (1 - IRR) \times 100$ . Effectiveness is reported only when the IRR was significantly less than 1.
d) Due to the absence of recorded events among 3-dose vaccinated children in these strata, the IRR was equal to zero, making it impossible to obtain adjusted estimates using the multivariable model. Therefore, we present the crude estimates restricted to these age strata and the 95% confidence intervals calculated using the exact Poisson method.

When evaluating incomplete series, the 1-dose series was significantly associated with an overall risk reduction of 38% (IRR 0.62, 95% CI 050–0.77). The 2-dose series was not statistically associated with the outcome (IRR 1.15, 95% CI 0.95–1.39). The effectiveness of these incomplete series was significantly modified by the child’s age (Supplemental Table S2), and we observed a significant benefit from the 1-dose and 2-dose series only in children under 1 year of age (IRR 0.43, 95% CI 0.26-0.71, and IRR 0.30, 95% CI 0.14-0.63, respectively; Table 3).

The 1- and 2-dose schedules of CoronaVac were associated with non-statistically significant reductions in the rate of COVID-19-attributed SARI in the 3–4-year age group compared with unvaccinated children (Table 4). Although no cases were identified among children who received a third dose of CoronaVac, this estimate was also not statistically significant (IRR 0; exact Poisson 95% CI 0–41.8). In age-specific analyses, the 2-dose schedule was associated with a higher rate of COVID-19-attributed SARI in the 3-year age group and with a non-statistically significant reduction in risk in the 4-year age group. Finally, when testing for interactions, we observed a positive and statistically significant interaction for the combination of one dose of CoronaVac and one dose of BNT162b2 (Supplemental Table S2).

**Table 4:** Incidence Rate Ratios of the CoronaVac vaccine against COVID-19-attributed SARI: overall and age-specific estimates for 3- and 4-year-old children.

|  | Adjusted IRR<br>(95 CI%)<sup>a</sup> | p-value |
| --- | --- | --- |
| **Overall estimate (3–4 years)** |  |  |
| 1 dose | 0.93 (0.54 - 1.62) | 0.81 |
| 2 doses | 0.96 (0.57 - 1.62) | 0.87 |
| 3 doses | 0.00 (0.00 - 41.78)<sup>b</sup> | 0.91 |
| **Age-specific estimate** |  |  |
| *3 years* |  |  |
| 1 dose | 1.07 (0.26 - 4.41) | 0.92 |
| 2 doses | 3.31 (1.31 - 8.37) | 0.01 |
| *4 years* |  |  |
| 1 dose | 0.97 (0.52 - 1.80) | 0.93 |
| 2 doses | 0.71 (0.37 - 1.36) | 0.30 |

Notes:
a) Adjusted for time and other vaccines (given the restriction to children aged 3–4 years, age was not included in the multivariable model)
b) Due to the absence of recorded events among 3-dose vaccinated children in these strata, the IRR was equal to zero, making it impossible to obtain adjusted estimates using the multivariable model. Therefore, we present the crude estimates restricted to these age strata and the 95% confidence intervals calculated using the exact Poisson method.

## DISCUSSION

In this population-based cohort study, we evaluated the first pediatric COVID-19 vaccines introduced in Brazil. Our results consistently indicate a high effectiveness of the complete 3-dose BNT162b2 primary series in reducing the COVID-19-attributed SARI rate in children aged 6 months to 4 years. However, fewer doses did not indicate consistent benefits, with effects being significantly modified by age.

The phase 2-3 clinical trial showed an efficacy of 73.2% (95% CI 43.8-87.6) for the 3-dose series of the BNT162b2 against symptomatic COVID-19 in this age group.^10^ However, as the authors noted, the trial did not have the power to assess efficacy against severe disease. Our estimates against SARI are higher, and this is consistent with current knowledge, as COVID-19 vaccines generally offer greater and more sustained protection against severe outcomes (such as hospitalizations) than against mild symptomatic infection. ^6,7^

Moving from efficacy to effectiveness, direct comparisons are limited since, to the best of our knowledge, no previous studies assessed the effectiveness of the complete 3-dose primary series of the BNT162b2 exclusively for children under 5 years of age. Existing systematic reviews primarily evaluate older pediatric groups (mean ages between 7 and 8 years) and focus on 1- or 2-dose series.^8^ Furthermore, our estimates in this age group also exceed the effectiveness observed with other mRNA vaccines. Specifically, a 2023 study in children aged 1 to 4 years reported that a complete 2-dose primary series (predominantly of mRNA-1273) exhibited a 63.3% (95% CI 40.6-77.3) effectiveness against omicron infection.^33^

Our age-specific analysis indicates that 1- or 2-dose schedules were beneficial only among children aged 6–11 months, with no observed benefit in those older than 1 year. This pattern aligns with clinical trial immunogenicity data showing that two doses elicited an insufficient immune response in children aged 2–4 years, supporting the need for a third dose.^10^

With CoronaVac, the reduction in COVID-19-attributed SARI risk in the 3–4-year age group was small and not statistically significant. Importantly, no deaths occurred among patients who received any dose of this vaccine. We cannot rule out the possibility that schedules with more doses can be associated with a greater benefit, but the third dose coverage in the studied population was insufficient to evaluate this hypothesis.

These results contrast with estimates from other populations, such as in Chile,^17^ where a study conducted in children aged 3 to 5 years indicated that a 2-dose CoronaVac series had an effectiveness of 64.6% (95% CI: 49.6–75.2) against COVID-19-associated hospitalizations (December 2021–February 2022). In part, this disagreement can be explained by differences in the observation period, since the Chilean study evaluated the protection soon after the vaccination, whereas we evaluated it over a longer and later period (July 2023-December 2024). Additionally, different outcome definitions (COVID-19-associated hospitalizations in Chile and stricter criteria for COVID-19-attributed SARI in our study) could contribute to the observed divergences.

As a limitation of this observational study, we cannot rule out unmeasured confounding, particularly related to determinants of vaccine uptake. For instance, among 1-year-old children, those who received one dose of CoronaVac had a higher hospitalization incidence than unvaccinated children (Table 2). Although this vaccine was not indicated for this age group (and therefore not included in our analysis), this finding may reflect preferential vaccination of children at higher baseline risk (e.g., severe comorbidities or immunosuppression). Such confounding by indication might similarly affect other evaluated schedules. Consequently, higher underlying risk among vaccinated children biases effectiveness toward the null. We adjusted for receipt of other vaccines, which may have partially mitigated this potential confounding (as suggested by our DAG).

Another potential limitation is the exclusive use of the campaign information system to identify the vaccinated cohort. At the end of the observation period, vaccinations were also recorded in a separate routine database, which was not linked by us due to the inability to identify and exclude duplicates. However, the high agreement with the official Ministry of Health dashboard (see Methods) suggests minimal underreporting. If present, such underreporting would underestimate vaccinated denominators, inflate their incidence rates, and bias effectiveness estimates towards the null, rendering our approach conservative.

Beyond the aforementioned potential biases that may have underestimated vaccine effectiveness, another relevant consideration is the natural immunity due to widespread viral circulation.^34–36^ In Brazil, the EPICOVID 2.0 study reported that 28.4% of the general population had experienced at least one COVID-19 infection by 2024, whereas a study conducted in India in June–July 2021 found a SARS-CoV-2 antibody prevalence of 68.3% among unvaccinated children.^36,37^ Thus, natural immunity among unvaccinated individuals could attenuate observed effectiveness estimates and may partly explain the absence of statistically significant associations for some vaccination schedules.

This phenomenon may also explain differences between epidemiological contexts. In Hong Kong,^18^ first dose BNT162b2 effectiveness among children aged 3 to 11 years was 65.6% (95% CI: 38.2–82.5), higher than our overall estimate (38%). That study occurred in early 2022 in a population with relatively low prior exposure to SARS-CoV-2. In contrast, our analysis took place later, when a higher prevalence of infection-acquired immunity would be expected, potentially reducing the relative benefit of vaccination.

Despite these challenges, the complete 3-dose BNT161b2 primary series maintained high and consistent effectiveness. We consider that this finding should be taken into account in the development of public health guidelines. Countries such as the United Kingdom,^19^ Norway^21^, and, recently in 2026, the United States, revised their universal routine vaccination recommendations, adopting a shared clinical decision-making approach.^20^ The main reason cited for this change was the shift in the cost-effectiveness ratio, considering the lower severity of the circulating variants and a higher prevalence of natural immunity, as discussed earlier.^36^ However, our results indicate that, in preventing severe COVID-19, routine vaccination for children aged 6 months to 4 years would reduce the healthcare system burden in scenarios where viral circulation remains endemic.

In conclusion, among the COVID-19 vaccination schedules introduced for children in Brazil, the complete 3-dose series of BNT162b2 demonstrated high and consistent effectiveness in preventing COVID-19–attributed SARI in children aged 6 months to 4 years. These findings support maintaining pediatric COVID-19 vaccination in the national routine schedule and strengthening efforts to ensure series completion.

## Data Availability

All data produced in the present study are available upon reasonable request to the authors. All the data sources are public and anonymized.

## SUPPLEMENTAL MATERIAL

**Figure S1:**
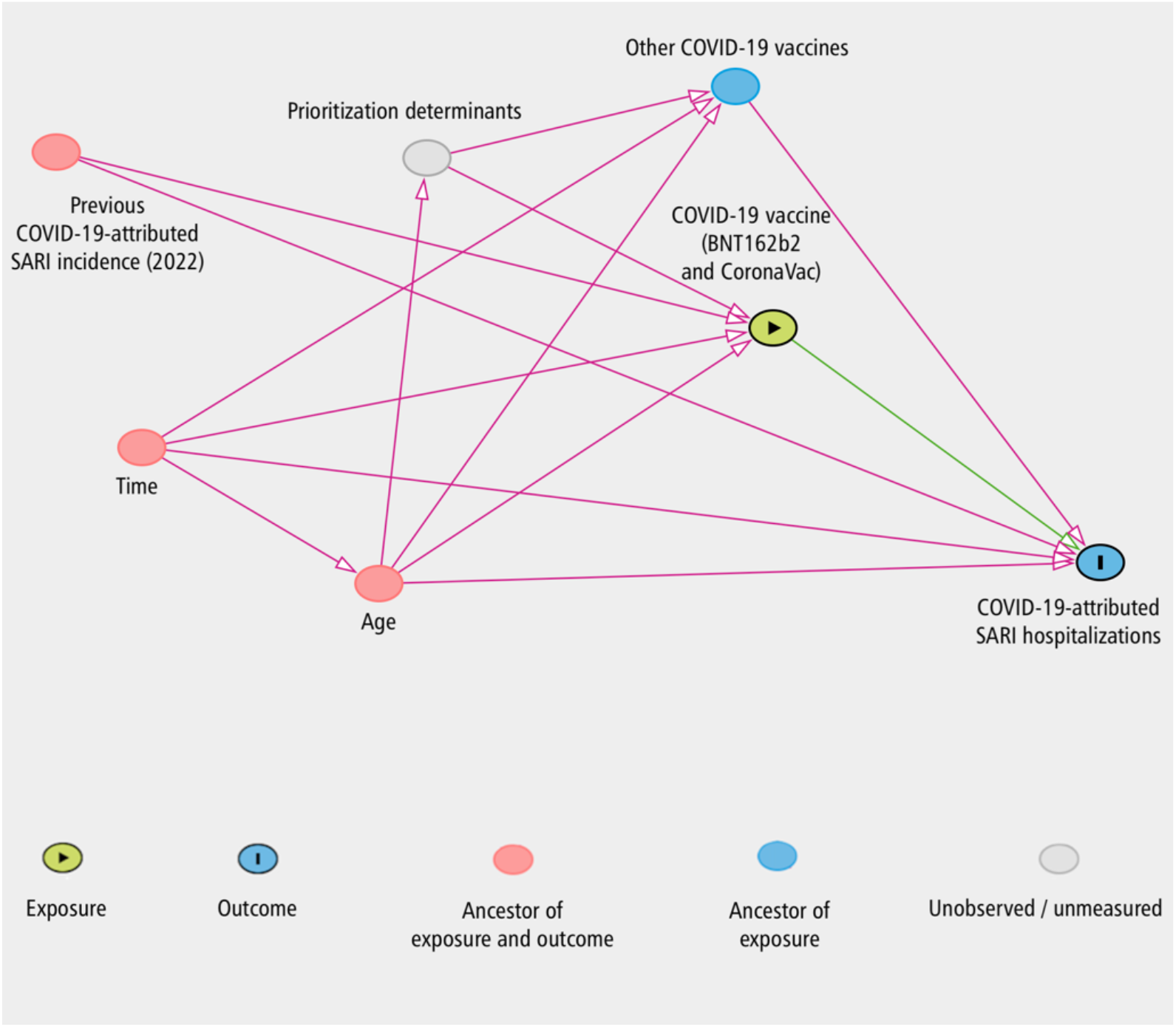
Directed acyclic graph (DAG) representing the causal relationship between the vaccine, COVID-19-attributed SARI hospitalizations and other covariates. Notes: The variables ‘Age’ and ‘Time’ were identified as common causes of the exposure and the outcome, justifying their inclusion in the model. The variable ‘Prioritization determinants’ represent unmeasured factors (such as comorbidities and healthcare access, among others) that influence both vaccination and SARI risk. The adjustment for ‘Other COVID-19 vaccines’ aims to partially block the confounding path generated by these unmeasured determinants.

**Table S1:**
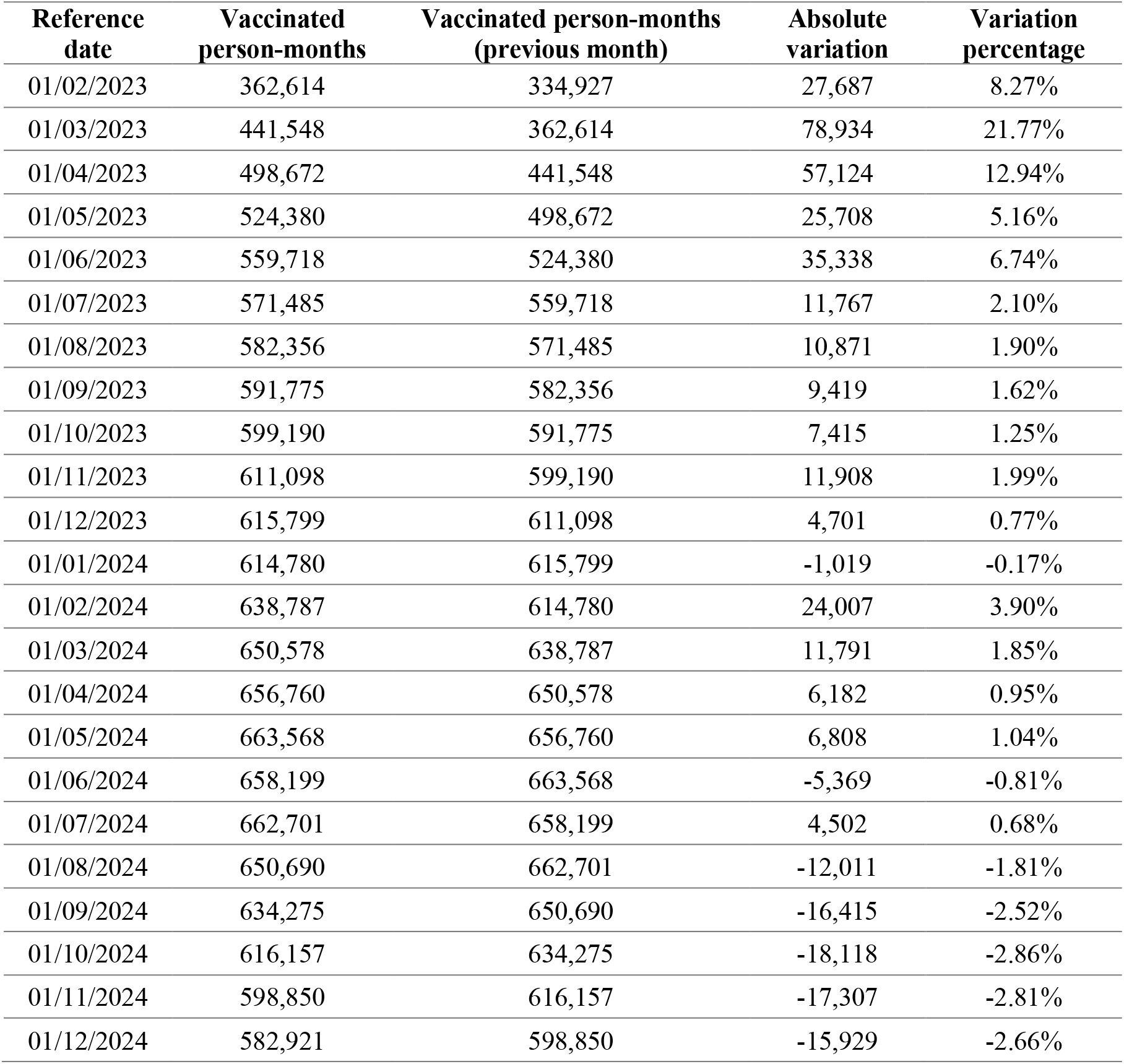
Absolute monthly variation and percentage in the populational denominator.

**Table S2:** Model with statistically significant effect modifiers (interactions) between vaccination status, age, and manufacturer combinations (July 2023-December 2024)

| Variable | Adjusted IRR (95 CI%) | p-value |
| --- | --- | --- |
| BNT162n2 doses (Ref: 0 doses) |  |  |
| 1 dose | 0.58 (0.46 - 0.74) | <0.001 |
| 2 doses | 0.80 (0.63 - 1.03) | 0.09 |
| 3 doses | 0.03 (0.01 - 0.10) | <0.001 |
| CoronaVac Doses (Ref: 0 doses) |  |  |
| 1 dose | 13.39 (3.71 - 48.35) | <0.001 |
| 2 doses | 2.76 (1.60 - 4.74) | <0.001 |
| Interactions |  |  |
| Age x 2 BNT162b2 doses | 1.73 (1.43 - 2.09) | <0.001 |
| Age x 1 CoronaVac dose | 0.44 (0.27 - 0.73) | < 0.001 |
| 1 BNT162b2 dose + 1 CoronaVac dose | 5.51 (2.07 - 14.69) | < 0.001 |

**Table S3:** Sensitivity analysis between the model with 24 municipalities and the model without the municipality of São Paulo due to the migration of its proprietary information system to the RNDS.

| Vaccine / Dose | Main model (24 municipalities) | Sensitivity (23 municipalities – without São Paulo) |
| --- | --- | --- |
| BNT162b2 - 3 Doses | IRR: 0.03 IC95%: 0.01 – 0.10<br>(p < 0.001) | IRR: 0.05 IC95%: 0.02 – 0.16<br>(p < 0.001) |
| CoronaVac - 2 Doses | IRR: 0.96 IC95%: 0.57 – 1.62<br>(p = 0.88) | IRR: 1.01 IC95%: 0.55 – 1.84<br>(p = 0.96) |

**Table S4:**
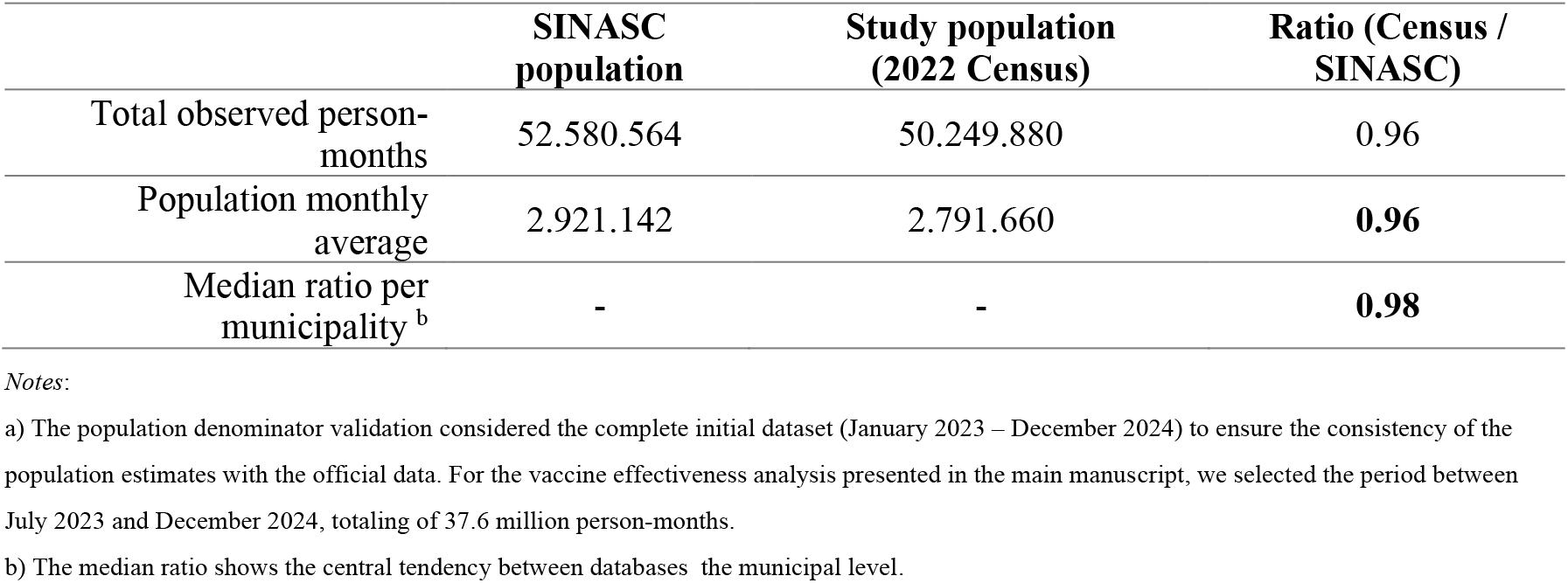
Comparison between the estimated population used in the study (based on the 2022 Census and vaccination records) and official live-birth records (SINASC) for the cohort aged 6 months to 4 years in the 24 selected municipalities, over the full 24 months of the database (January 2023-December 2024) ^a^.

|  | SINASC population | Study population (2022 Census) | Ratio (Census / SINASC) |
| --- | --- | --- | --- |
| Total observed person-months | 52.580.564 | 50.249.880 | 0.96 |
| Population monthly average | 2.921.142 | 2.791.660 | **0.96** |
| Median ratio per municipality <sup>b</sup> | - | - | **0.98** |

Notes:
- a) The population denominator validation considered the complete initial dataset (January 2023 – December 2024) to ensure the consistency of the population estimates with the official data. For the vaccine effectiveness analysis presented in the main manuscript, we selected the period between July 2023 and December 2024, totaling of 37.6 million person-months. - b) The median ratio shows the central tendency between databases the municipal level.

**Table S5:**
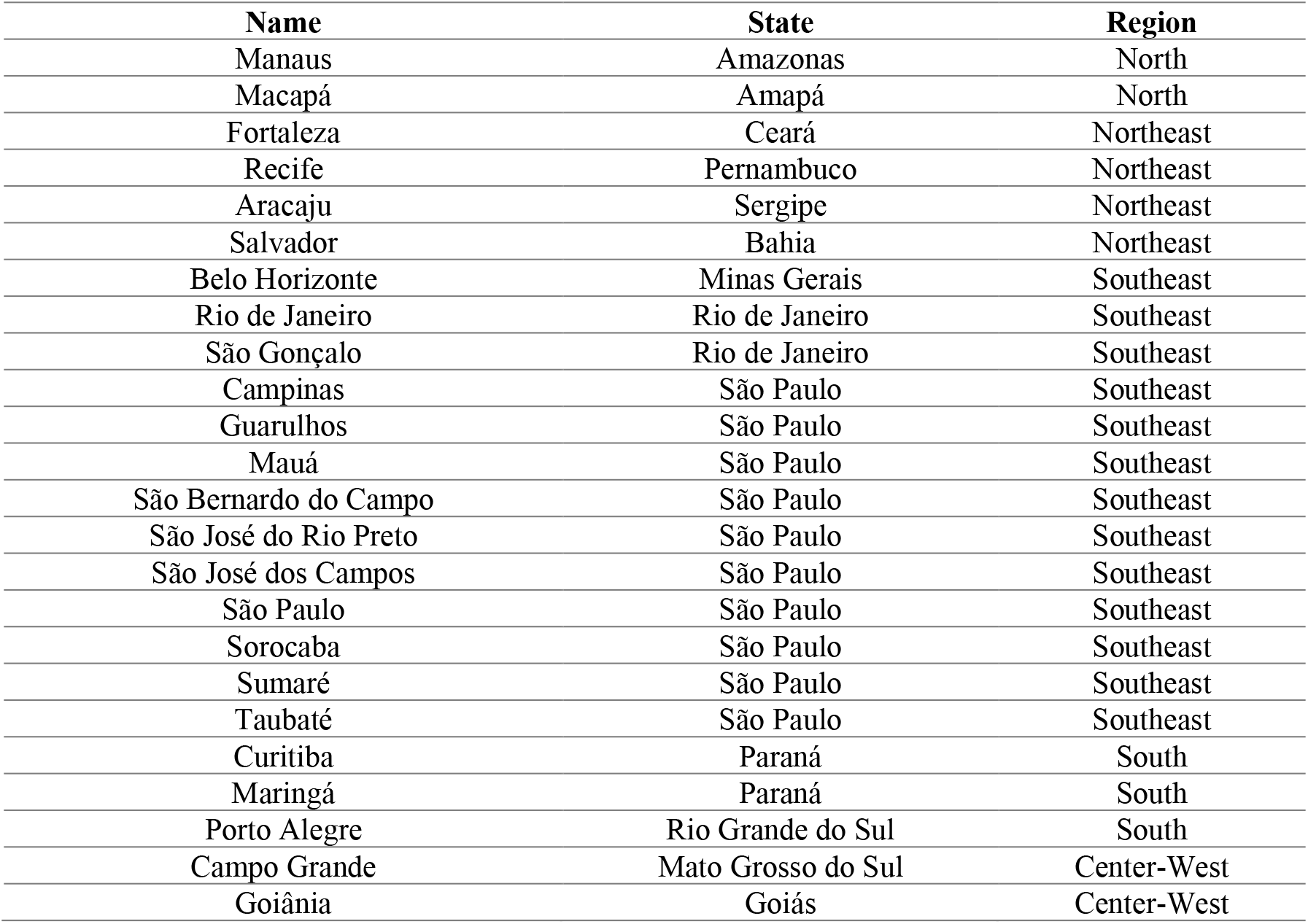
List with the 24 municipalities used in the study, with name, State and Region.

## REFERENCES

1. Leite IS, Ribeiro DAG, Vieira ILV, Gama FO da. A evolução das coberturas vacinais brasileiras e os impactos provocados pela pandemia de Covid-19 nas metas de imunização. Research, Society and Development 2022;11(11):e205111133041– e205111133041.

2. Domingues CMAS, Teixeira AM da S, Moraes JC de. Vaccination coverage in children in the period before and during the COVID-19 pandemic in Brazil: a time series analysis and literature review. Jornal de Pediatria 2023;99:S12–21.

3. Procianoy GS, Rossini Junior F, Lied AF, Jung LFPP, Souza MCSC de. Impact of the COVID-19 pandemic on the vaccination of children 12 months of age and under: an ecological study. Ciênc saúde coletiva 2022;27:969–78.

4. Sato APS. Pandemia e coberturas vacinais: desafios para o retorno às escolas. Rev Saúde Pública 2020;54:115.

5. Soares MCB, Freitas BAC de, Toledo LV, Mendes IR, Quintão AP de C, Souza SM de. Hospitalizations and deaths of children and adolescents with Severe Acute Respiratory Infection due to COVID-19 during the epidemiological year of 2020. Rev Inst Med Trop Sao Paulo 2023;65:e11.

6. Chi W-Y, Li Y-D, Huang H-C, et al. COVID-19 vaccine update: vaccine effectiveness, SARS-CoV-2 variants, boosters, adverse effects, and immune correlates of protection. J Biomed Sci 2022;29(1):82.

7. Andrews N, Stowe J, Kirsebom F, et al. Covid-19 Vaccine Effectiveness against the Omicron (B.1.1.529) Variant. N Engl J Med 2022;386(16):1532–46.

8. Hu Y-H, Ding Y, Li L-W, Cheng J, Cai Y-H. Evaluation of vaccine effectiveness of mRNA COVID-19 vaccines in children: a systematic review and meta-analysis. Eur Rev Med Pharmacol Sci 2024;28(6):2584–92.

9. Anderson EJ, Campbell JD, Creech CB, et al. Warp Speed for Coronavirus Disease 2019 (COVID-19) Vaccines: Why Are Children Stuck in Neutral? Clin Infect Dis 2021;73(2):336–40.

10. Muñoz FM, Sher LD, Sabharwal C, et al. Evaluation of BNT162b2 Covid-19 Vaccine in Children Younger than 5 Years of Age. New England Journal of Medicine 2023;388(7):621–34.

11. Scutari R, Fox V, Nguyen JL, et al. Clinical manifestations and severity of COVID-19 caused by Omicron among paediatric patients aged 0–17 years in Italy. Sci Rep 2025;15(1):36638.

12. Saito S, Asai Y, Matsunaga N, et al. First and second COVID-19 waves in Japan: A comparison of disease severity and characteristics. J Infect 2021;82(4):84–123.

13. Costa VC da, Montarroyos UR, Lopes KA de M, Santos ACOD. A Series of Severe and Critical COVID-19 Cases in Hospitalized, Unvaccinated Children: Clinical Findings and Hospital Care. Epidemiologia (Basel) 2025;6(3):40.

14. Ministério da Saúde, Brasil. NOTA TÉCNICA N° 399/2022-CGPNI/DEIDT/SVS/MS - Recomendação da vacina COVID-19 Pfizer-BioNTech para crianças de 6 meses a 4 anos de idade (4 anos, 11 meses e 29 dias). [Internet]. Secretaria de Vigilância em Saúde e Ambiente (SVSA); 2022 [cited 2026 Jan 22]. Available from: https://www.gov.br/saude/pt-br/centrais-de-conteudo/publicacoes/notas-tecnicas/2022/nota-tecnica-no-399-2022-cgpni-deidt-svs-ms

15. Ministério da Saúde, Brasil. NOTA TÉCNICA N° 213/2022-CGPNI/DEIDT/SVS/MS [Internet]. Secretaria de Vigilância em Saúde e Ambiente (SVSA); 2022. Available from: https://www.gov.br/saude/pt-br/centrais-de-conteudo/publicacoes/notas-tecnicas/2022/nota-tecnica-213-2022-cgpni-deidt-svs-ms

16. Vacinometro COVID-19 [Internet]. 2025 [cited 2025 Apr 4]; Available from: https://infoms.saude.gov.br/extensions/SEIDIGI_DEMAS_Vacina_C19/SEIDIGI_DEMAS_Vacina_C19.html

17. Jara A, Undurraga EA, Zubizarreta JR, et al. Effectiveness of CoronaVac in children 3–5 years of age during the SARS-CoV-2 Omicron outbreak in Chile. Nat Med 2022;28(7):1377–80.

18. Rosa Duque JS, Leung D, Yip KM, et al. COVID-19 vaccines versus pediatric hospitalization. Cell Rep Med 2023;4(2):100936.

19. JCVI statement on COVID-19 vaccination in 2025 and spring 2026 [Internet]. GOV.UK. [cited 2026 Jan 24]; Available from: https://www.gov.uk/government/publications/covid-19-vaccination-in-2025-and-spring-2026-jcvi-advice/jcvi-statement-on-covid-19-vaccination-in-2025-and-spring-2026

20. US Department of Health and Human Services. Fact Sheet: CDC Childhood Immunization Recommendations [Internet]. 2026 [cited 2026 Jan 24]; Available from: https://www.hhs.gov/press-room/fact-sheet-cdc-childhood-immunization-recommendations.html

21. Coronavirus vaccine for children 5-11 years [Internet]. Norwegian Institute of Public Health. 2022 [cited 2026 Jan 24]; Available from: https://www.fhi.no/en/archive/covid-19-archive/koronavaksine---arkiverte-artikler/coronavirus-vaccine-for-children-5-11-years/

22. Ladhani SN. COVID-19 vaccination for children aged 5–11 years. The Lancet 2022;400(10346):74–6.

23. Ministério da Saúde, Brasil. Guia - Vigilância integrada da covid-19, influenza e outros vírus respiratórios de importância em saúde pública [Internet]. [cited 2026 Jan 24]; Available from: https://www.gov.br/saude/pt-br/centrais-de-conteudo/publicacoes/guias-e-manuais/2024/guia-vigilancia-integrada-da-covid-19-influenza-e-outros-virus-respiratorios-de-importancia-em-saude-publica/view

24. Brasil. Portal de Dados Abertos [Internet]. 2024 [cited 2024 July 25]; Available from: https://dados.gov.br/dados/conjuntos-dados/covid-19-vacinacao1

25. Ministério da Saúde, Brasil. PORTARIA GM/MS N° 5.201, DE 15 DE AGOSTO DE 2024 - Lista nacional de notificação compulsória de doenças, agravos e eventos de saúde pública [Internet]. [cited 2026 Jan 24]; Available from: https://bvsms.saude.gov.br/bvs/saudelegis/gm/2024/prt5201_19_08_2024.html

26. Ministério da Saúde. RNDS - Rede Nacional de Dados em Saúde [Internet]. 2024 [cited 2024 Jan 9]; Available from: https://www.gov.br/saude/pt-br/composicao/seidigi/rnds

27. Falcão MZ, Rachid R, Fornazin M. AI innovation in healthcare and state platforms under a rights-based perspective: the case of Brazillian RNDS. Data & Policy 2024;6:e70.

28. População residente, por sexo, idade e forma de declaração da idade - Censo 2022 [Internet]. [cited 2024 Sept 9]; Available from: https://sidra.ibge.gov.br/tabela/9514

29. Firely. Rede Nacional de Dados em Saúde - RNDS | Grupo de Atendimento - SIMPLIFIER.NET [Internet]. 2025 [cited 2025 Apr 23]; Available from: https://simplifier.net/redenacionaldedadosemsaude/codesystem-brgrupoatendimento

30. Diaz-Quijano FA, Carvalho DS de, Raboni SM, et al. Effectiveness of mass dengue vaccination with CYD-TDV (Dengvaxia®) in the state of Paraná, Brazil: integrating case-cohort and case-control designs. The Lancet Regional Health – Americas [Internet] 2024 [cited 2024 July 26];35. Available from: https://www.thelancet.com/journals/lanam/article/PIIS2667-193X(24)00104-2/fulltext

31. Ministério da Saúde, Brasil. PORTARIA GM/MS N° 5.663, DE 31 DE outubro DE 2024 - DOU - Imprensa Nacional [Internet]. [cited 2026 Jan 24]; Available from: https://www.in.gov.br/web/dou

32. Ministério da Saúde, Brasil. Sistema de Informações sobre Nascidos Vivos (SINASC) - CGIAE - DAENT - SVSA/MS [Internet]. 2024 [cited 2024 July 25]; Available from: https://svs.aids.gov.br/daent/cgiae/sinasc/

33. Wee LE, Tang N, Pang D, et al. Effectiveness of Monovalent mRNA Vaccines Against Omicron XBB Infection in Singaporean Children Younger Than 5 Years. JAMA Pediatr 2023;177(12):1324–31.

34. Clarke KEN, Jones JM, Deng Y, et al. Seroprevalence of Infection-Induced SARS-CoV-2 Antibodies - United States, September 2021-February 2022. MMWR Morb Mortal Wkly Rep 2022;71(17):606–8.

35. Naeimi R, Sepidarkish M, Mollalo A, et al. SARS-CoV-2 seroprevalence in children worldwide: A systematic review and meta-analysis. EClinicalMedicine 2023;56:101786.

36. Ghosh A, Goyal K, Singh R, et al. High prevalence of severe acute respiratory syndrome coronavirus-2 (SARS-CoV-2) antibodies among unvaccinated children of Chandigarh, Northwest India, in a household-based paediatric serosurvey post-second wave of pandemic (June to July 2021). Public Health 2023;225:160–7.

37. Wehrmeister FC, Menezes AMB, Dalcolmo M, et al. A countrywide study on post-COVID-19 condition in Brazil: the Epicovid 2.0. Int J Epidemiol 2025;55(Suppl 1):i31–40.

